# Late diagnosis of infants with PCD and neonatal respiratory distress

**DOI:** 10.1101/2020.03.27.20045377

**Authors:** Goutaki Myrofora, S Halbeisen Florian, Barbato Angelo, Crowley Suzanne, Harris Amanda, A Hirst Robert, Karadag Bülent, Martinu Vendula, Morgan Lucy, O’Callaghan Chris, Ozçelik Ugur, Scigliano Sergio, Ucros Santiago, Yiallouros Panayiotis, M Schulzke Sven, E Kuehni Claudia

## Abstract

Neonatal respiratory distress (NRD) is common among infants with primary ciliary dyskinesia (PCD), but we do not know whether affected neonates are diagnosed timely.

We used data from the international PCD cohort study (iPCD), including only participants diagnosed between 2000 and 2019 using current diagnostic criteria. We assessed the proportion of patients with PCD with a history of NRD and their age at diagnosis, stratifying by presence of laterality defects. First we analysed data from the entire cohort and then from a subgroup including children diagnosed using stricter criteria.

Among the 1375 patients in the study, 45% had a history of NRD and 42% a laterality defect. Out of the 476 children with definite PCD diagnosis, 55% had a history of NRD and 50% a laterality defect. PCD was diagnosed at a median age of 3.4 years in this group, varying from less than 1 year in Norway and Cyprus to 10 years in Turkey. Overall, 30% of children with PCD were diagnosed during the first 12 months of life. This varied from 13% in those with situs solitus and no NRD, to 21% in those with situs solitus and NRD, 33% in those with situs anomalies but no NRD, and 52% in those with both NRD and situs anomalies.

Our results suggest that we need to improve our knowledge of the neonatal presentation of infants with PCD, and apply this knowledge in neonatology so that these patients will receive appropriate care sooner, at the start of their extrauterine life.

**What is already known on this topic?:**

- Many patients with primary ciliary dyskinesia (PCD) present with neonatal respiratory distress (NRD).
- The neonatal period would therefore be an ideal window of opportunity to diagnose PCD early, before long-term damage to the lungs has occurred.
- Despite this, PCD is usually diagnosed late in life.

**What this study adds?:**

- 55% of children with PCD in this large multinational dataset had a history of NRD.
- Among these, PCD was diagnosed early in those with laterality defects (median age 0.9 years) but late in those with situs solitus (5.9 years).
- This suggests that neonatologists and paediatricians do not suspect PCD as a cause of NRD in term infants unless it is accompanied by laterality defects.

## Introduction

Primary ciliary dyskinesia (PCD) is a multiorgan genetic disease that affects approximately 1 in 10,000 people ^1 2^, or as many as 1 in 400 in highly consanguineous populations. ^3 4^ The clinical phenotype is variable, but most patients have chronic upper and lower airway disease with rhinitis and cough resulting in recurrent infections of ears, sinuses, and lungs.^5 6^ About half have situs inversus, and an additional 10-12% other laterality defects, which may be associated with congenital heart disease.^7 8^ Already in childhood, lung function is comparable to patients with cystic fibrosis (CF), and as the disease progresses most patients develop progressive lung disease with bronchiectasis and chronic pseudomonas infection.^9-13^ At advanced disease stages, many adults become oxygen dependent, undergo lobectomy and some will eventually need lung transplantation.^14 15^ It is believed that as in other genetic respiratory diseases such as CF, early diagnosis followed by initiation of regular physiotherapy and prompt antibiotic treatment may reduce lung function decline and improve long-term outcomes.^16^

Patients with PCD are usually born at term, and typically present with chronic respiratory symptoms from birth, mainly rhinitis and a wet-sounding cough. A large proportion present with neonatal respiratory distress (NRD).^17^ The frequency of those symptoms in neonates with PCD is unclear; in a systematic review, the proportion of PCD patients reported to have NRD varied widely between studies, from 15% to 91%. However, this data was generally of poor quality.^18^ In a single-centre case-control study from Canada, detailed neonatal data were extracted from health records to identify characteristics that distinguish PCD from other causes of NRD.^19^ The study showed that compared to other term infants with NRD, infants with PCD had a characteristic clinical picture that should point toward the diagnosis.

## Objective

The neonatal period offers an ideal window for early diagnosis of PCD before long-term damage to the lungs has occurred. However, the practice of timely diagnosis has not yet been investigated in an international setting. We used a multicentre dataset from the international PCD (iPCD) cohort to determine the proportion of patients presenting with NRD and the age when PCD was finally diagnosed. We grouped infants into those with and without NRD, and stratified additionally by presence of laterality defects.

## Methods

### Participants

The iPCD cohort, described in detail elsewhere, included 3824 patients as of June 2019 when reviewed for this study.^20^ The core dataset of the iPCD cohort is compulsory for all contributors, while specific modules including the one on neonatal history are voluntary. For this analysis, we included only patients from data providers who had also contributed data on the neonatal period for their patients (Figure 1 and supplementary Table S1). We further excluded patients diagnosed before the year 2000 because PCD diagnosis has evolved considerably during the past 20 years and because we were mainly interested in recent data, representative of contemporary neonatal care.^21^

**Fig 1.**
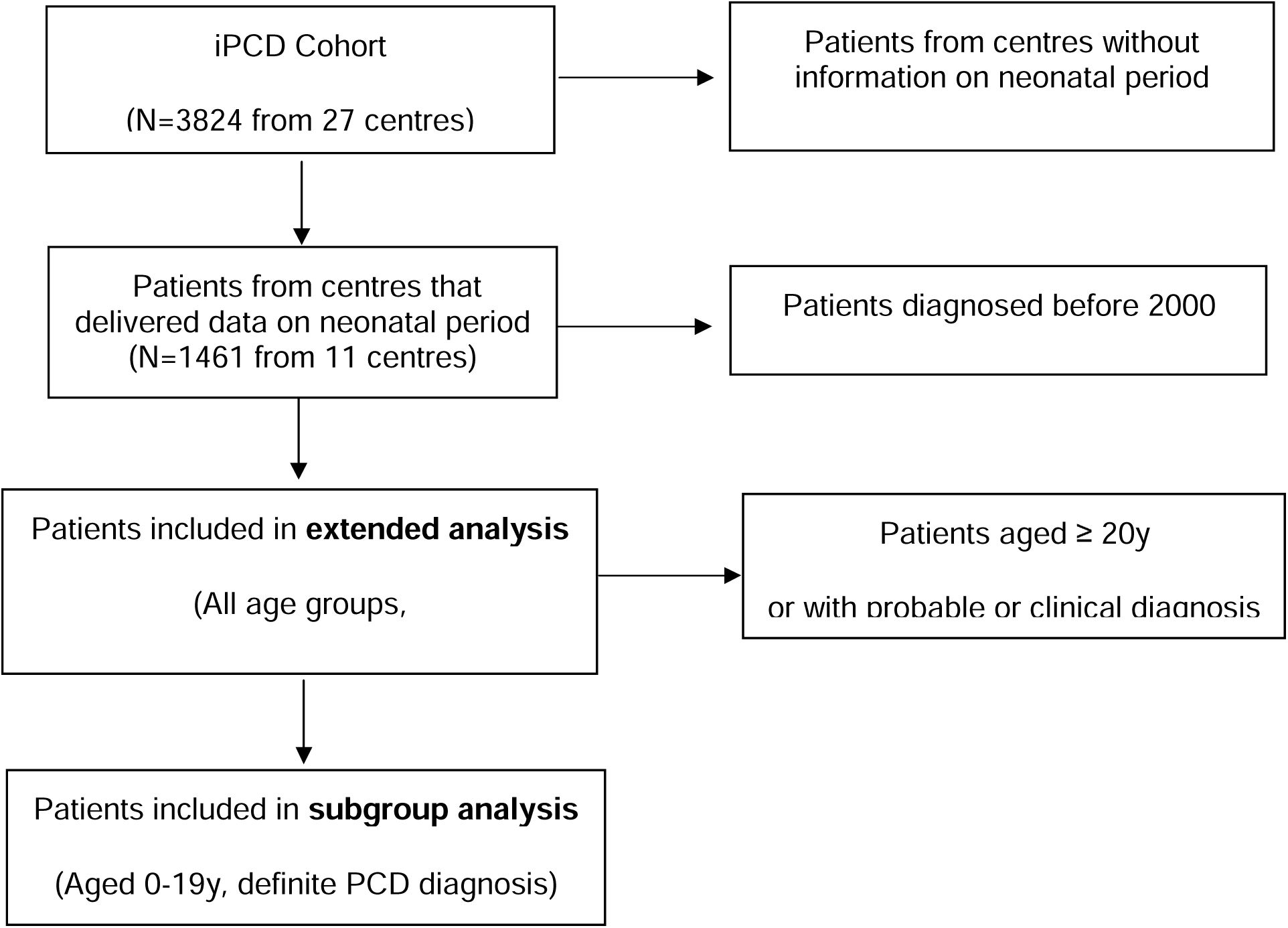
Flow chart showing the patients included for the different analyses performed. NRD: neonatal respiratory distress.

### Definition of NRD and PCD diagnosis

NRD was described as a history of respiratory distress during the neonatal period as reported by either patients or parents, or retrieved from hospital records. In one of the centres, NRD was defined as presence of respiratory distress or physician-recorded chest symptoms during the neonatal period since the data did not distinguish between the two. We also retrieved information on laterality status and defined the presence of situs inversus or heterotaxia as laterality defect. All patients included in the study had a clinical phenotype consistent with PCD and were under PCD management at the contributing centres.

Because the diagnosis of PCD is complex and can sometimes remain inconclusive we grouped patients into groups of diagnostic certainty based on the diagnostic guidelines of the European Respiratory Society Task Force.^22^

“Definite PCD” diagnosis was defined as a pathogenic biallelic PCD genetic mutation or hallmark structural abnormality in transmission electron microscopy (TEM).^22^ The remaining patients had either abnormal high-speed video microscopy findings or low nasal nitric oxide (probable PCD diagnosis), or were patients with strong clinical suspicion in whom other possible diagnoses such as CF were excluded, but the diagnostic algorithm was not complete (clinical PCD diagnosis). Age at diagnosis was defined based on information provided by the centres or, when not available, based on the date of positive diagnostic tests. For patients with clinical diagnosis, age of diagnosis was defined as the age when the patient started to be managed as PCD at the participating centre. We divided the age at diagnosis into periods, to investigate whether the diagnosis was made in the first three months of life, within infancy (age 3-11 months), in the preschool period (age 1-4 years) or during school years (age 5-9, 10-14 or over 14 years).

### Analysis

We first analysed data from all patients regardless of age or diagnostic certainty (Figure 1), and assessed the proportion of patients who reported NRD and laterality defects, overall and by country. We then stratified the population into four groups and described their age at diagnosis: patients without NRD and with situs solitus, patients with NRD and situs solitus, patients without NRD and with laterality defect and patients with both NRD and laterality defect. We assessed the proportion of children diagnosed with PCD within the first three and the first 12 months of life, and the proportion diagnosed later at ages 1-4, 5-9, 10-14, or 15-19 years. We also compared age at diagnosis using Kruskal-Wallis test, followed by pairwise comparisons between groups using a Wilcoxon rank sum test.

Second, we repeated all analyses, in patients who had “definite PCD” based on recent diagnostic guidelines and were younger than 20 years at the time of investigation, because recall of neonatal problems declines with age (subgroupanalysis, Figure 1).^22 23^ We used STATA 15.1 (StataCorp, College Station, TX, USA) for all analyses. While the first dataset is representative for the majority of patients with PCD currently in treatment, the second includes those for whom quality of information on PCD diagnosis and neonatal period is best.

### Ethics

In most participating countries, researchers are not required to obtain patient informed consent for retrospectively collected anonymised observational data. In countries where informed consent was needed, PIs obtained locally ethics approval and informed consent for the contribution of their anonymised data to the iPCD cohort for research purposes.^20^

## Results

Out of the 3824 patients included in the iPCD cohort at the time of the study, 1461 originated from centres that completed also the module on the neonatal period (Figure 1). After excluding patients diagnosed earlier than 2000, 1375 patients were included in the study (extended analysis). The subgroup analysis included 476 patients who were 0-19 years old and had a definite PCD diagnosis (Figure 1). Data came from 11 centres in nine countries: Argentina, Australia, Colombia, Cyprus, Czech Republic, Italy, Norway, Turkey, and the United Kingdom (UK). Seven countries were represented by one centre, Turkey and UK had two contributing centres each.

In the entire study population (N=1375), 45% (95%CI 43-48%) had a history of NRD and 42% (95%CI 40-45%) a laterality defect (Supplementary Table S2). PCD was diagnosed at a median age of 9.8 years. Only 13% of the 1375 were diagnosed during the first 12 months of life, varying from 4% in those with situs solitus and no NRD to 32% in those with NRD and laterality defect (Table S2). Median age at diagnosis was 12.4 years for patients without NRD and situs solitus, 10.4 years for those with NRD and situs solitus, 8.8 years for those without NRD but with a laterality defect and 4.5 years for the group combining a history of NRD and laterality defect. In 14% of patients with NRD and a laterality defect, PCD was diagnosed in the first three months of life (Supplementary Table S3). Among patients with NRD but without laterality defect, PCD was only diagnosed in the first three months of life in 2.9%.

Results were comparable in the subgroup analyses that included only patients 0-19 years with definite PCD diagnosis. 55% (95%CI 50-59%) reported a history of NRD and 50% (95%CI 46-55%) had a laterality defect (Table 1). Prevalence of NRD and laterality defects varied between centres (Supplementary Table S4). Age at diagnosis overall was lower than in the whole study population and the differences between the four groups more pronounced. Patients were diagnosed at a median age of 3.4 years, varying from less than one year in Norway and Cyprus to ten years in Turkey (Table 1). Overall, 30% of children with PCD were diagnosed during the first 12 months of life, varying from 13% in those with no NRD and situs sollitus, to 21% in those with NRD and situs solitus, 33% in those no NRD and laterality defect, and 52% in those with both NRD and laterality defect.

**Table 1.** Characteristics of PCD patients aged 0-19 years with definite PCD diagnosis (N=476)

| **Characteristic** | **N (%)** | **Age at diagnosis**<br>**(in years)**<br>**Median (IQR)** | **N (%) of patients**<br>**diagnosed**<br>**at 0-12m** |
| --- | --- | --- | --- |
| **Total** | 476 (100) | 3.36 (0.69-7.76) | 144 (30.3) |
| **Sex** |  |  |  |
| Male | 262 (55.0) | 2.62 (0.61-7.06) | 87 (33.2) |
| Female | 214 (45.0) | 4.38 (0.97-8.34) | 57 (26.6) |
| **Country of residence** |  |  |  |
| Argentina | 30 (6.3) | 1.5 (0.50-5.00) | 10 (33.3) |
| Australia | 19 (4.0) | 1.26 (0.40-3.78) | 9 (47.4) |
| Colombia | 11 (2.3) | 6.61 (4.04-8.34) | 1 (9.1) |
| Cyprus | 5 (1.0) | 0.61 (0.21-0.67) | 4 (80.0) |
| Czech Republic | 21 (4.4) | 4.75 (1.49-9.28) | 4 (19.0) |
| Italy | 89 (18.7) | 1.62 (0.38-5.67) | 36 (40.4) |
| Norway | 25 (5.3) | 0.63 (0.10-7.17) | 14 (56.0) |
| Turkey | 31 (6.5) | 10.11 (7.37-12.53) | 0 (0.0) |
| United Kingdom | 245 (51.5) | 3.53 (0.89-8.08) | 66 (26.9) |
| **NRD** |  |  |  |
| No | 193 (40.6) | 4.96 (1.10-8.44) | 43 (22.3) |
| Yes | 261 (54.8) | 1.96 (0.50-6.40) | 98 (37.5) |
| No information | 22 (4.6) | 2.47 (1.42-9.40) | 3 (13.6) |
| **Organ laterality** |  |  |  |
| Situs solitus | 218 (45.8) | 5.93 (2.00-9.08) | 37(17.0) |
| Laterality defect | 240 (50.4) | 1.44 (0.42-5.07) | 103 (42.9) |
| No information | 18 (3.8) | 6.35 (1.09-9.54) | 4 (22.2) |
| **Clinical characteristics groups** |  |  |  |
| No NRD, situs solitus | 114 (24.0) | 6.47 (2.73-9.08) | 15 (13.2) |
| NRD, situs solitus | 132 (27.7) | 5.44 (1.50-9.41) | 27 (20.5) |
| No NRD, laterality defect | 91 (19.1) | 2.51 (0.67-6.97) | 30 (33.0) |
| NRD, laterality defect | 139 (29.2) | 0.91 (0.31-3.17) | 72 (52.0) |

NRD: neonatal respiratory distress

Table 2 and Figure 2 show a detailed breakdown of age at diagnosis for the four patient groups. For infants with NRD and a laterality defect, median age at diagnosis was 0.9 years: 21% were diagnosed within three months of birth, a further 31% before the age of 12 months, and most (80%) before they reached five years. Children with laterality defects not presenting with neonatal respiratory symptoms were also diagnosed relatively early (median age 2.5 years): one-third were diagnosed during infancy and 57% before the age of five years. These results stand in contrast with those for infants with situs solitus. Among infants who presented with NRD and situs solitus, only 6% were diagnosed with PCD within three months, 20% within the first year of life, and only 39% by age five. Of the children, who had situs solitus, PCD was diagnosed at a median age of 5.4 years if they had NRD, but of 6.5 years if they had no neonatal symptoms (p=0.15, Figure 2), suggesting that neonatal symptoms alone, in the absence of situs anomalies, do not lead to diagnostic testing for PCD.

**Table 2.** Age at diagnosis of PCD patients aged 0-19 years with definite PCD diagnosis, per clinical characteristics group, based on reported NRD and organ laterality (N=476)

| Clinical characteristics groups | Age at diagnosis N (%) |  |  |  |  |  | TOTAL |
| --- | --- | --- | --- | --- | --- | --- | --- |
|  | 0-2 m | 3-11 m | 1-4 yrs | 5-9 yrs | 10-14 yrs | >14 yrs |  |
| No NRD, situs solitus | 2 (1.7) | 13 (11.4) | 24 (21.1) | 46 (40.4) | 26 (22.8) | 3 (2.6) | 114 (100) |
| NRD, situs solitus | 8 (6.1) | 19 (14.4) | 25 (18.9) | 45 (34.1) | 34 (25.7) | 1 (0.8) | 132 (100) |
| No NRD, laterality defect | 6 (6.6) | 24 (26.3) | 22 (24.2) | 27 (29.7) | 11 (12.1) | 1 (1.1) | 91 (100) |
| NRD, laterality defect | 29 (20.9) | 43 (30.9) | 39 (28.1) | 20 (14.4) | 7 (5.0) | 1 (0.7) | 139 (100) |
| TOTAL | 45 (9.4) | 99 (20.8) | 110 (23.1) | 138 (29.0) | 78 (16.4) | 6 (1.3) | 476 (100) |

NRD: neonatal respiratory distress, m: months, yrs: years
Age 0-2 months corresponds to 0-2.999 months, similar for all age groups.

**Fig 2.**
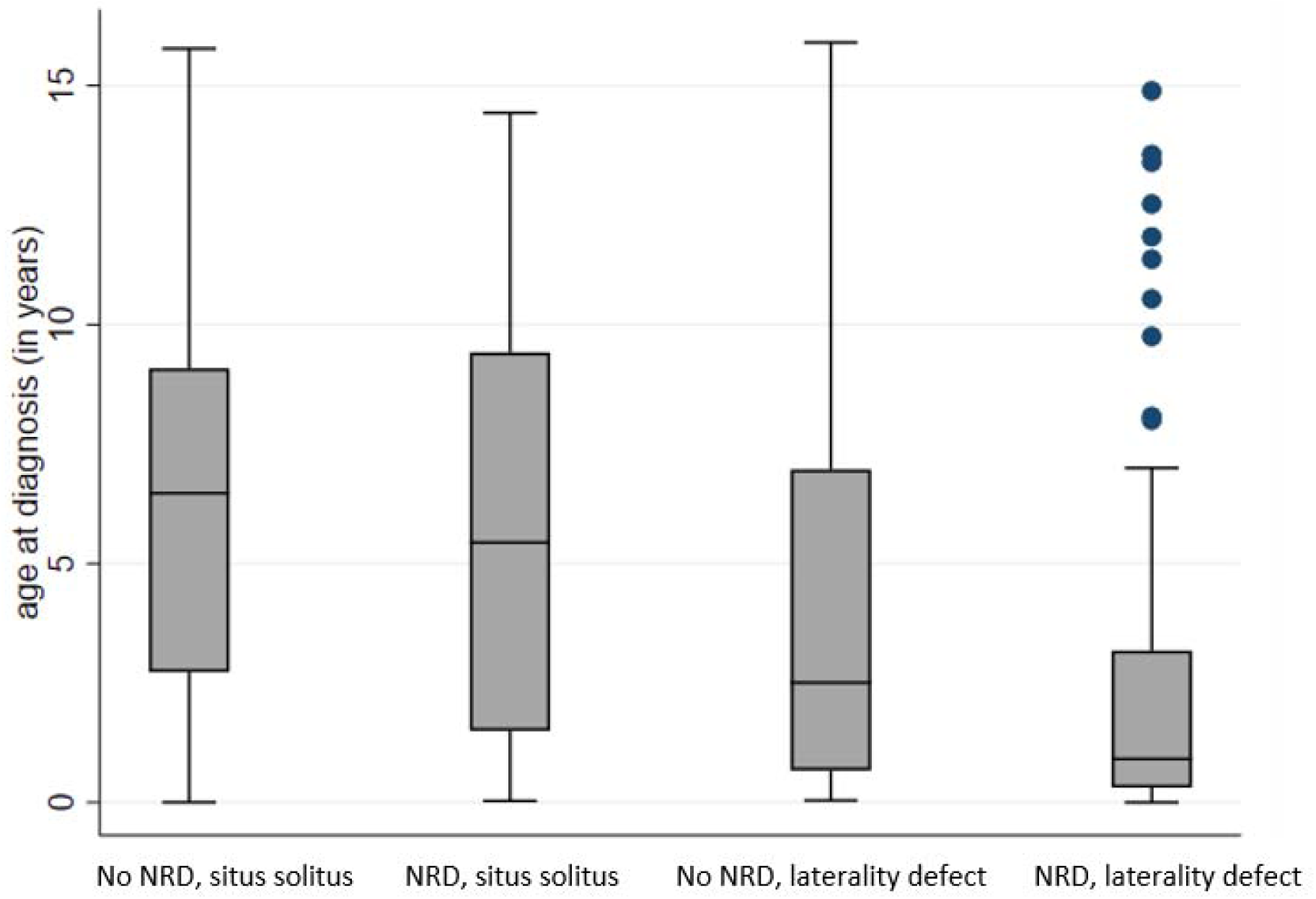
Age at diagnosis of PCD patients (N=476) aged 0-19 years with definite PCD diagnosis, per clinical characteristics group; NRD: neonatal respiratory distress.

Each box represents the median and interquartile range (IQR) of age at diagnosis for the respective group (in years). The whiskers represent the range of age at diagnosis and dots represent outliers;

Pairwise comparisons between the four groups using a Wilcoxon rank sum test corrected for multiple testing (Benjamini-Hochberg) resulted in p≤0.003, with the exemption of the comparison between infants with no NRD and situs solitus with infants with NRD and situs solitus (p=0.150)

## Discussion

This is the first large multinational study that describes the proportion of infants with PCD who presented with respiratory distress after birth, and the age they were diagnosed. Among the 55% of children with PCD in the iPCD cohort who reported NRD, those who also had situs inversus were diagnosed with PCD relatively early, at a median age of 0.9 years, while those who had no laterality defect were diagnosed at a median age of 6 years, independently of whether or not they had presented with NRD. This suggests that although NRD in infants with PCD has typical features, the diagnosis is usually missed by neonatologists and paediatricians unless patients also present with situs anomalies.

The iPCD cohort is the largest dataset on patients with PCD worldwide, and is representative of currently diagnosed and followed PCD patients in developed countries.^20^. We analysed the whole dataset, which was large and representative for the majority of PCD patients in medical follow-up, and then, to reduce uncertainties in diagnosis of PCD and of recall bias regarding the neonatal presentation, we repeated all analyses for patients with a definite diagnosis of PCD based on current diagnostic criteria, including only those who were younger than 20 when neonatal history was taken. Still, our study has limitations. In particular, we lacked detailed information on the characteristics of the neonatal disease presentation, such as onset and duration of respiratory distress, exact diagnoses and treatments given at the hospital and results of x-rays and other investigations. Thus, we cannot state whether the clinical picture of NRD was as typical as that previously described. ^19^

There are few other studies to compare our findings with. Prevalence of NRD in our study was lower than in a single-centre study from Canada, which was based on a chart review of neonatal records (91%).^19^ In our study, neonatal history was usually obtained from the patients or the parents at the time of PCD diagnosis. NRD prevalence varied between countries and was low in some centres. In several centres the prevalence of NRD was higher (e.g. 68-100%), suggesting that the recorded average might be an underestimation due to a less detailed history in some centres. In another study of consecutively referred and diagnosed patients with PCD from the UK, 56 of 75 patients (75%) reported neonatal chest symptoms.^17^ As in our study, these data were also collected from patients or their parents at the first diagnostic visit. The prevalence found was higher than ours (45%). A possible explanation for this difference might be that the definition used for NRD in the UK study was wider as it included also other neonatal chest symptoms while in our study it included only history of NRD (with the exception of one centre).

What implication does this have for clinical management? Given that up to 5% of term-born infants present with some type of respiratory distress and assuming that 50% of the infants with PCD present with NRD, and estimating prevalence of PCD as 1 in 10000, it follows that around 1 in 500 neonates with NRD has PCD.^24^ Performing the complex and expensive set of diagnostic tests to diagnose one infant with PCD in 500 is unrealistic, if not impossible. However, the data from the Canadian case-control study suggest (though this needs to be confirmed in other populations) that the clinical picture of NRD is quite typical in PCD, and it differs substantially from much more common diagnoses such as transient tachypnea of the newborn or peripartum pneumothorax given that infants with PCD often required supplemental oxygen for several days to weeks and had lobar collapse. In that study, the combination of oxygen therapy for more than two days and lobar collapse in the chest x-ray had a sensitivity of 83% for detecting PCD.^19^ Thus, term-born infants with late onset but long duration of respiratory distress not needing intubation, and with a radiological picture typical for PCD could be picked up as suspected PCD and referred for diagnosis from the first days of life. In our study, the average age at diagnosis for children in some centres e.g. in Norway and Cyprus was 7 to 8 months, suggesting, although the number of patients was low, that earlier diagnosis is possible.

## Conclusions

In analogy to CF, and based on the (comparatively) sparse evidence available in PCD, it is thought that early introduction of specialised physiotherapy, a meticulous vaccination scheme, prompt use of antibiotic therapy in respiratory infections and prophylactically if required, avoidance of active and passive tobacco smoking, and nutritional support when needed has a positive impact on the trajectory of lung function, general health, quality of life, and the lifespan of PCD patients.^16 25-27^ We found that only a minority of infants with PCD who present typical symptoms in the neonatal period are diagnosed early, but examples from some countries suggest this is possible. In term neonates with NRD, better recognition of the typical clinical picture associated with PCD in the neonatal period could increase the proportion of neonates referred for detailed PCD diagnostic testing. We believe that if the typical clinical picture of PCD in neonates can be better characterised and its recognition is improved in the neonatal period, more infants suffering from PCD could receive appropriate care, sooner.

## Data Availability

The ethical approval of the study does not allow to have the datasetopenly available. Data are available to researchers upon request by the study PIs

## Authors’ contributions

MG and CK developed the concept and designed the study. MG and FH cleaned and standardised the data, and performed the statistical analyses. All other authors participated in discussions for the development of the study and contributed data. MG and CK drafted the manuscript; all authors contributed to iterations and approved the final version. MG and CK take final responsibility for the contents.

## Acknowledgements

We thank all the patients with PCD in the cohort and their families, and especially the PCD patient organisations for their close collaboration. We also thank all the researchers in the participating centres who were involved in data collection and data entry, and worked closely with us through the whole process of participating to the iPCD Cohort. We thank Christopher Ritter (Institute of Social and Preventive Medicine, University of Bern, Switzerland) for his editorial suggestions to this manuscript.

